# *APOE* Genotype-specific Methylation Patterns are Linked to Alzheimer Disease Pathology and Estrogen Response

**DOI:** 10.1101/2022.12.20.22283744

**Authors:** Rebecca Panitch, Nathan Sahelijo, Junming Hu, the Alzheimer’s Disease Neuroimaging Initiative, Kwangsik Nho, David A. Bennett, Kathryn L. Lunetta, Rhoda Au, Thor D. Stein, Lindsay A. Farrer, Gyungah R. Jun

## Abstract

The joint effects of *APOE* genotype and DNA methylation on Alzheimer disease (AD) risk is relatively unknown. We conducted genome-wide methylation analyses using 2,021 samples in blood (91 AD cases, 329 mild cognitive impairment, 1,391 controls) and 697 samples in brain (417 AD cases, 280 controls). We identified differentially methylated levels in AD compared to controls in an *APOE* genotype-specific manner at 25 cytosine-phosphate-guanine (CpG) sites in brain and 36 CpG sites in blood. Additionally, we identified seven CpG sites in the *APOE* region containing *TOMM40, APOE*, and *APOC1* genes with P<5×10^−8^ between *APOE* ε4 carriers and non-carriers in brain or blood. In brain, the most significant CpG site hypomethylated in ε4 carriers compared to non-carriers) was from the *TOMM40* in the total sample, while most of the evidence was derived from AD cases. However, the CpG site was not significantly modulating expression of these three genes in brain. Three CpG sites from the *APOE* were hypermethylated in *APOE* ε4 carriers in brain or blood compared in ε4 non-carriers and nominally significant with *APOE* expression in brain. Three CpG sites from the *APOC1* were hypermethylated in blood, which one of the 3 CpG sites significantly lowered *APOC1* expression in blood using all subjects or ε4 non-carriers. Co-methylation network analysis in blood and brain detected eight methylation networks associated with AD and *APOE* ε4 status. Five of the eight networks included genes containing network CpGs that were significantly enriched for estradiol perturbation, where four of the five networks were enriched for the estrogen response pathway. Our findings provide further evidence of the role of *APOE* genotype on methylation levels associated with AD, especially linked to estrogen response pathway.

## Introduction

Alzheimer disease (AD) is a neurodegenerative disorder characterized neuropathologically by neurofibrillary tangles and amyloid plaques [1]. The apolipoprotein E (*APOE*) ε4 variant is the strongest genetic risk factor for late-onset AD, while the ε2 variant has been shown to confer protection against AD, in a dose-dependent manner [2,3,4]. Single copies of the ε4 and ε2 alleles are associated with 3 to 4-fold increased and 0.61-fold decreased risk of AD, respectively. Previous studies identified *APOE* genotype-specific mechanisms including the complement pathway and blood-brain barrier dysfunction [5,6,7].

Large-scale genome-wide association studies (GWAS) have identified contributions to AD risk from more than 75 independent loci, but the large portion of heritability of the disease is unexplained [8]. Emerging omics technologies have prompted investigations of gene expression and epigenetic profiles at the tissue and cellular levels. For example, it has been shown that the degree of methylation of cytosine-phosphate-guanine (CpG) dinucleotides in brain differ between AD cases and controls in novel regions as well as in loci previously associated with AD risk such as *BIN1* [9]. Methylation levels at multiple CpG sites assessed in peripheral blood have also been associated with cognitive decline and AD progression [10].

Several CpG sites in the *APOE* region are differentially methylated in AD cases compared to controls, and distinct methylation patterns have been observed between persons with the ε3/ε3 and ε3/ε4 genotypes [11]. In addition, the *APOE* region has been shown to be differentially methylated between healthy ε2 and ε4 carriers in blood [12]. However, despite these findings, the effect of *APOE* genotypes, especially on the genome-wide level for AD risk remains relatively unknown. Here, we analyzed methylation array data from blood and brain tissue in three datasets to discover *APOE* genotype-dependent genome-wide associations of methylation with AD risk and related traits, as well as co-methylation networks.

## Methods

### Sources of Methylation and Phenotypic Data

Data were obtained for participants of three cohort studies including the Religious Orders Study and Rush Memory and Aging Project (ROSMAP), Alzheimer’s Disease Neuroimaging Initiative (ADNI), and Framingham Heart Study (FHS).

#### Religious Orders Study and Rush Memory and Aging Project (ROSMAP)

Clinical, neuropathological information, *APOE* genotyping, and preprocessed, quality controlled, and normalized brain HumanMethylation450 BeadChip methylation array data derived from dorsolateral prefrontal cortex area tissue donated by 697 ROSMAP participants (417 autopsy-confirmed AD cases and 280 controls) [13,14,15] were obtained from the CommonMind Consortium portal (http://www.synapse.org) (**Supplementary Table 1**). AD diagnosis was determined using National Institute of Aging (NIA) Reagan criteria for intermediate or high probability of AD [16]. AD-related traits included Braak staging for neurofibrillary tangles [17] and the Consortium to Establish a Registry for Alzheimer Disease (CERAD) semi-quantitative criteria for measuring neuritic plaques (CERAD Score) [18]. ROS and MAP were both approved by an Institutional Review Board of Rush University Medical Center. All participants signed an informed consent, Anatomic Gift Act, and repository consent.

#### Alzheimer’s Disease Neuroimaging Initiative (ADNI)

Infinium® MethylationEPIC BeadChip beta values and phenotype data were obtained from the LONI website (http://adni.loni.usc.edu) for 630 ADNI participants including 91 with a clinical AD diagnosis, 329 with mild cognitive impairment (MCI), and 210 controls [10]. Methylation array iDAT files were processed and normalized using wateRmelon [19]. Because methylation was measured in DNA extracted from blood specimens obtained at multiple examinations, methylation data from the earliest timepoint were analyzed. Among the available endophenotypic data, analyses included magnetic resonance imaging (MRI) volumetric measures of ventricles, and hippocampus, and entorhinal thickness, as well as neuropsychological test scores consisted of the Alzheimer’s Disease Assessment Scale – 13 item (ADAS13), Clinical Dementia Rating Scale Sum of Boxes (CDRSB), logical memory - delayed recall (LDELTOTAL), Rey Auditory Verbal Learning Test (RAVLT) immediate, RAVLT learning, and RAVLT percent forgetting.

#### Framingham Heart Study (FHS)

Cognitive test and normalized HumanMethylation450 BeadChip methylation array data were obtained for 1,391 cognitively healthy participants from the generation 3 cohort in FHS [20]. Cognitive test scores at the same time point of methylation measurement included the Paired Associate Learning - Recognition (PASr) test, Logical Memories – Immediate Recall (LMi) test, Logical Memories – Delayed Recall (LMd) test, Similarities Test (SIM), Visual Reproductions – Delayed Recall (VRd), Trail A (trailsA) test, the animal portion of the Verbal Fluency Test (FAS_animal), and Boston Naming Test (BNT30).

### Differential Methylation Analysis

Differential methylation between AD and control samples was performed in the ADNI and ROSMAP datasets using the LIMMA software [21]. The methylation percentage of each CpG site, defined as the proportion of total signal from the methylation-specific probe, was compared between AD cases and controls using linear regression models including sex, age, and batch as covariates. Genome-wide methylation analyses were conducted in the total sample and separately within *APOE* ε4 carriers (ε2/ε4, ε3/ε4, and ε4/ε4) and non-carriers (ε2/ε2, ε2/ε3, and ε3/ε3). Genome-wide methylation analysis between *APOE* ε4 carriers and non-carriers was performed in the ADNI and ROSMAP datasets using LIMMA and regression models including terms for age, sex and batch in the total sample and separately within AD and control groups.

Genome-wide methylation analysis between *APOE* ε4 carriers and non-carriers for each CpG site was conducted in FHS using lmekin package in R and a linear mixed effect model incorporating a genetic relatedness matrix (GRM) as a random effect with sex and age at exam as covariates. The GRM was generated using genetic dosage data and the software GCTA [22] to account for familial relationships among 8,481 FHS participants.

### Association of Methylation with Expression of Genes in the *APOE* Region

RNA-sequencing (RNA-seq) data derived from ROSMAP brains were obtained and processed as previously described [7]. Matched RNA-seq and methylation data were available for 510 ROSMAP participants (297 AD cases, 213 controls). Normalized gene-expression microarray data were obtained from the LONI website (http://adni.loni.usc.edu) for 159 ADNI subjects (42 AD cases, 117 controls) who also had matching methylation array data. Significantly methylated CpG sites between *APOE* ε4-carriers and non-carriers (p=5.0×10^−8^) in *APOE* region genes (*NECTIN2, APOC1, APOE*, and *TOMM40*) were selected for further analysis. The association of gene expression level and methylation levels at the CpG sites in the *APOE* region was evaluated using linear regression models with covariates including age, sex, RNA integrity number (RIN), RNA batch, and methylation batch. Post-mortem interval (PMI) was included as an additional covariate in analyses of the ROSMAP dataset that had this information. Analyses were performed in the total sample and separately within *APOE* ε4 carriers and non-carriers.

### Association of Methylation with Phenotypic Traits

Quantitative or semi-quantitative traits were rank-transformed after adjusting for age and sex as previously described [23]. In the ROSMAP dataset, the association of CpG site methylation with rank-transformed Braak stage and CERAD score was assessed using regression models including batch as a covariate. In the ADNI dataset, the association of CpG site methylation with cognitive test scores and imaging phenotypes were assessed using regression models including covariates of batch and education for cognitive traits and of batch and intracranial volume for imaging phenotypes. In the FHS dataset, we tested the association of methylation with cognitive test scores using a linear mixed effects model accounting for education and family structure with the GRM as covariates. Association models were evaluated in the total sample and separately within groups of *APOE* ε4 carriers and non-carriers.

### Co-methylation Network Analysis

Co-methylation networks were generated with differentially methylated CpG sites (P<0.05) between *APOE* ε4 carriers and non-carriers in the ADNI and ROSMAP datasets using the weighted correlation network analysis (WGCNA) program [24]. Analyses in the ADNI dataset also included data from 329 subjects with MCI. We selected four and six soft-power parameters in the ADNI and ROSMAP datasets, respectively, as previously described [5]. CpG percentages were hierarchically clustered using a dissimilatory topological overlap matrix (TOM). Modules with a minimum of 100 CpG sites were created using dynamic tree cutting, networks with similar eigenvalues and a height of 0 were merged using WGCNA’s mergeCloseModules function. The signedKME function assigned fuzzy module membership. We identified networks exhibiting significantly different methylation levels of eigenvalues between *APOE* ε4 carriers and non-carriers and between AD cases and controls, determined by a student’s t-test, which were selected for subsequent analysis. Biological pathways (MSigDB_Hallmark_2020) and drug perturbations (Drug_Perturbations_from_GEO_2014) for each network were identified using the EnrichR program applied to genes containing CpG sites in significant networks [25]. QIAGEN Ingenuity Pathway Analysis (IPA) software was used to create a biological network containing overlapping genes across modules from the EnrichR analysis.

## Results

### Differentially methylated CpG sites between AD cases and controls

Methylation levels in 697 brain samples from ROSMAP participants and 301 blood samples from ADNI participants were compared between AD cases and controls in the total sample, and within *APOE* ε4 carrier and non-carrier subgroups (**Figure 1a and Supplementary Table 1**). In the combined datasets, there were no genome-wide significant (P<5×10^−8^) differentially methylated CpG sites between AD cases and controls in those with or without ε4. However, analysis of the brain data revealed, moderately different (P<10^−5^) methylation levels at 3 CpG sites among ε4 carriers and 22 CpG sites among ε4 non-carriers (**Table 1** and **Supplementary Figure 1a**). Of the 25 CpG sites that were differentially methylated in either *APOE* genotype subgroup, approximately half (13 CpG sites) were moderately differentially methylated in the total sample. Most of the CpG sites that were differentially methylated among ε4 non-carriers (20/25 = 80%) were significantly associated (P<2.0×10^−3^) with Braak stage and/or CERAD score (**Figure 1b and Supplementary Table 2**). Methylation levels of two intergenic CpG sites (cg05731218 and cg12307200) were lower in AD cases compared to controls in the total sample and associated with both Braak stage and CERAD score at a genome-wide significance level (P<5×10^−8^). The most significant association of methylation CpG sites located within genes were observed for cg10907744 in *GPR133* with Braak stage (P=5.8×10^−6^) and CERAD score (P=4.6×10^−6^) and for cg19987111 in *CHSY1* with Braak stage (P=8.5×10^−8^).

**Table 1.** Differentially methylated CpG sites in brain for AD in total, *APOE* ε4 carriers, and non-carriers

| CpG Name | Chr | Position | Gene | Total Sample | | *APOE* ε4 carriers | | *APOE* ε4 non-carriers | |
| --- | --- | --- | --- | --- | --- | --- | --- | --- | --- |
|  |  |  |  | T | P | T | P | T | P |
| cg19533050 | 2 | 163175044 | IFIH1 | -1.23 | 0.22 | -4.92 | $1.9 \times 10^{-6}$ | 0.35 | 0.73 |
| cg23808213 | 2 | 166948291 | SCN1A | -4.81 | $1.9 \times 10^{-6}$ | -1.17 | 0.24 | -4.71 | $3.2 \times 10^{-6}$ |
| cg05731218 | 2 | 216769199 | intergenic | -7.06 | $4.2 \times 10^{-12}$ | -2.97 | $3.3 \times 10^{-3}$ | -5.15 | $3.7 \times 10^{-7}$ |
| cg04436449 | 3 | 185214835 | TMEM41A | -4.43 | $1.1 \times 10^{-5}$ | -0.56 | 0.57 | -4.62 | $4.9 \times 10^{-6}$ |
| cg12307200 | 3 | 188664632 | intergenic | -7.04 | $4.7 \times 10^{-12}$ | -4.48 | $1.3 \times 10^{-5}$ | -4.87 | $1.5 \times 10^{-6}$ |
| cg16234490 | 4 | 77138082 | FAM47E | -4.48 | $8.6 \times 10^{-6}$ | 0.17 | 0.87 | -4.97 | $9.2 \times 10^{-7}$ |
| cg24899806 | 7 | 119914282 | KCND2 | -3.77 | $1.8 \times 10^{-4}$ | 0.22 | 0.83 | -4.51 | $8.2 \times 10^{-6}$ |
| cg23831517 | 8 | 34182528 | intergenic | 4.25 | $2.5 \times 10^{-5}$ | -0.63 | 0.53 | 4.63 | $4.5 \times 10^{-6}$ |
| cg14096074 | 9 | 34255149 | KIF24 | -4.40 | $1.3 \times 10^{-5}$ | -0.99 | 0.32 | -4.66 | $4.0 \times 10^{-6}$ |
| cg03727169 | 10 | 31418969 | intergenic | -4.67 | $3.6 \times 10^{-6}$ | -1.60 | 0.11 | -4.94 | $1.1 \times 10^{-6}$ |
| cg01982597 | 10 | 50733420 | ERCC6 | -4.10 | $4.7 \times 10^{-5}$ | 0.54 | 0.59 | -4.64 | $4.4 \times 10^{-6}$ |
| cg20326704 | 10 | 70321770 | TET1 | -1.57 | 0.12 | -4.61 | $7.6 \times 10^{-6}$ | 0.27 | 0.79 |
| cg04126866 | 10 | 85932763 | C10orf99 | -5.24 | $2.2 \times 10^{-7}$ | -1.84 | 0.07 | -4.59 | $5.5 \times 10^{-6}$ |
| cg14882481 | 11 | 107437051 | ALKBH8 | -4.54 | $6.7 \times 10^{-6}$ | 0.19 | 0.85 | -4.89 | $1.4 \times 10^{-6}$ |
| cg10907744 | 12 | 131589455 | GPR133 | 4.97 | $8.6 \times 10^{-7}$ | 2.56 | 0.01 | 4.56 | $6.4 \times 10^{-6}$ |
| cg18708502 | 13 | 21588555 | LATS2 | 4.19 | $3.1 \times 10^{-5}$ | -0.18 | 0.86 | 4.50 | $8.4 \times 10^{-6}$ |
| cg16746221 | 14 | 20666088 | OR11G2 | -3.86 | $1.2 \times 10^{-4}$ | -1.15 | 0.25 | -4.49 | $8.7 \times 10^{-6}$ |
| cg24231804 | 15 | 67316861 | intergenic | -5.53 | $4.5 \times 10^{-8}$ | -1.73 | 0.09 | -4.68 | $3.7 \times 10^{-6}$ |
| cg14829066 | 15 | 88559141 | NTRK3 | -4.66 | $3.8 \times 10^{-6}$ | -1.06 | 0.29 | -5.15 | $3.7 \times 10^{-7}$ |
| cg19987111 | 15 | 101747167 | CHSY1 | 4.10 | $4.6 \times 10^{-5}$ | -1.65 | 0.10 | 4.75 | $2.7 \times 10^{-6}$ |
| cg02432274 | 16 | 88378468 | intergenic | 2.00 | 0.05 | 4.68 | $5.5 \times 10^{-6}$ | 0.02 | 0.98 |
| cg05952786 | 17 | 48559485 | RSAD1 | -5.11 | $4.2 \times 10^{-7}$ | -1.67 | 0.10 | -4.87 | $1.5 \times 10^{-6}$ |
| cg15503752 | 17 | 74639731 | ST6GALNAC1 | 4.40 | $1.2 \times 10^{-5}$ | -0.52 | 0.60 | 4.62 | $4.9 \times 10^{-6}$ |
| cg05421550 | 19 | 4446485 | UBXN6 | -4.88 | $1.3 \times 10^{-6}$ | -1.66 | 0.10 | -5.11 | $4.6 \times 10^{-7}$ |
| cg19612770 | 19 | 4475216 | HDGF2 | -4.82 | $1.7 \times 10^{-6}$ | -1.49 | 0.14 | -4.73 | $2.9 \times 10^{-6}$ |

Only CpG sites moderately ( $P < 10^{-5}$ ) differentially methylated in either *APOE* ε4 carriers and/or non-carriers were included.
T: T-value, P: P-value

**Figure 1:**
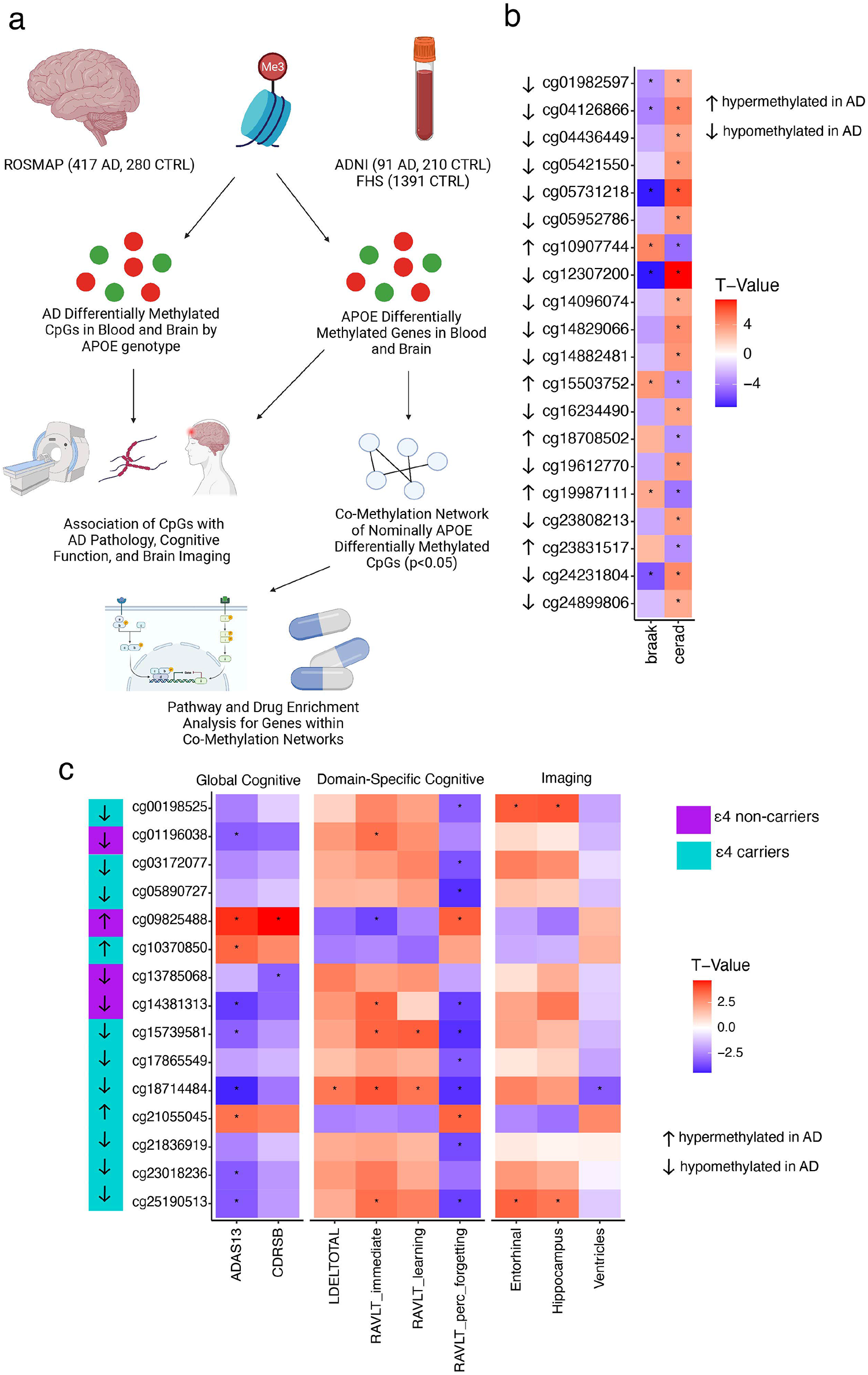
Differential methylation between AD cases and controls grouped by *APOE* ε4 carrier status. **(a)** Study Design. Methylation array data were obtained from blood (2 datasets) and frozen brain tissue (1 dataset). Methylation of CpG sites was compared between AD cases and controls as well as between *APOE* ε4 carriers and non-carriers. Association of methylation at CpG sites with neuropathological, cognitive and imaging traits was also evaluated. CpG sites with nominally significant P values (P<0.05) between *APOE* ε4 carriers and non-carriers were incorporated in co-methylation network analyses performed separately for data derived from blood and brain. Finally, biological pathways and drug perturbations were identified from analyses of co-methylation networks. Figure created with biorender.com. **(b)** Heatmap showing association of neuropathological traits with methylation at CpG sites that were differentially methylated (P<10^−5^) between AD cases and controls. CpG sites whose degree of methylation was significantly (P<2.0×10^−3^) associated with multiple testing correction with at least one trait are indicated by an asterisk. **(c)** Heatmap showing association of cognitive and MRI imaging traits with methylation at CpG sites that were differentially methylated (P<10^−5^) between AD cases and controls. Direction of differential methylation between AD and controls in *APOE* ε4 carriers or non-carriers in blood was shown. CpG sites whose degree of methylation was significantly associated with at least one trait after multiple testing correction (P<1.4×10^−3^) are indicated by an asterisk.

Moderately significant differential methylation between AD cases and controls from blood were observed in 21 CpG sites among ε4 carriers and 15 CpG sites in ε4 non-carriers (**Supplementary Table 3 and Supplementary Figure 1b**). In contrast to the findings in the brain data, none of these CpG sites were improved in significance in the total sample (**Supplementary Table 3**). Methylation of the 15 CpG sites, eleven among ε4 carriers and four among ε4 non-carriers, was significantly (multiple testing correction P<1.4×10^−3^) associated with performance on global and domain-specific cognitive tests, and/or MRI brain imaging measures (**Figure 1c**). Methylation of the CpG site cg09825488 in *EXO5* was increased in AD cases compared to controls in ε4 non-carriers (P=2.9×10^−6^, **Supplementary Table 3**) and significantly associated with both global cognitive tests and several domain specific cognitive tests (**Supplementary Table 4**). Methylation of three CpG sites (cg00198525, cg18714484, and cg25190513) in ε4 carriers was significantly associated with the volume of cortical brain regions (**Supplementary Table 4**). Methylation at the cg18714484 in *CHEK1* was decreased in AD cases compared to controls in ε4 carriers (P=2.2×10^−6^; **Supplementary Table 3**) and inversely associated with global cognitive (P=1.8×10^−5^), memory performance (4.2×10^−5^) and ventricle volume (P=8.1×10^−4^).

### Differentially methylated CpG sites between *APOE* ε4 carriers and non-carriers

Eight CpG sites were significantly differentially methylated (5×10^−8^) between *APOE* ε4 carriers and non-carriers in the ADNI, FHS, and ROSMAP datasets, and seven of the 8 sites are located within the *APOE* region (chr19:45380000-45430000) (**Table 2 and Figure 2a**). In brain, cg02613937 located in *TOMM40* was the most significant CpG site (hypomethylated in ε4 carriers compared to non-carriers) in the total sample (P=1.3×10^−13^) and most of the evidence was derived from AD cases (P=7.0×10^−13^). Methylation at the cg02613937 was not significantly associated with expression of genes in the *APOE* region (**Figure 2b**). In contrast, three CpG sites from the *APOE* were hypermethylated in *APOE* ε4 carriers in brain (cg14123992, cg04406254) and blood (cg06750524) compared in ε4 non-carriers **(Table 2, Figure 2a**). Methylation at both *APOE* CpG sites in brain was nominally associated (P<0.05) with the *APOE* expression in ε4 carriers only (**Figure 2b**). Three CpG sites from the *APOC1* in blood were significantly hypomethylated in ε4 carriers compared to non-carriers in either AD cases (cg07773593) or controls (cg23270113 and cg05644480). Methylation of cg07773593 was nominally significant (p<0.05) with lower *APOC1* expression in the total sample and ε4 non-carriers (**Supplementary Figure 2**).

**Table 2.** Differentially methylated CpG sites in brain for *APOE* □4 status in total, AD cases, and controls

| CpG Name | Chr | Position | Source | Gene | Total Sample |  | AD |  | Control |  |
| --- | --- | --- | --- | --- | --- | --- | --- | --- | --- | --- |
|  |  |  |  |  | T | P | T | P | T | P |
| cg05002071 | 11 | 76510323 | FHS (blood) | *intergenic* | 5.67 | $1.5 \times 10^{-8}$ | NA | NA | 5.67 | $1.5 \times 10^{-8}$ |
| cg02613937 | 19 | 45395297 | ROSMAP (brain) | *TOMM40* | -7.55 | $1.3 \times 10^{-13}$ | -7.41 | $7.0 \times 10^{-13}$ | -3.26 | $1.3 \times 10^{-3}$ |
| cg14123992 | 19 | 45407868 | ROSMAP (brain) | *APOE* | 6.74 | $3.4 \times 10^{-11}$ | 6.51 | $2.2 \times 10^{-10}$ | 2.93 | $3.7 \times 10^{-3}$ |
| cg04406254 | 19 | 45407945 | ROSMAP (brain) | *APOE* | 6.05 | $2.4 \times 10^{-9}$ | 5.90 | $7.4 \times 10^{-9}$ | 2.45 | 0.02 |
| cg06750524 | 19 | 45409955 | FHS (blood) | *APOE* | 6.45 | $1.1 \times 10^{-10}$ | NA | NA | 6.45 | $1.1 \times 10^{-10}$ |
| cg23270113 | 19 | 45417587 | FHS (blood) | *APOC1* | -6.40 | $1.5 \times 10^{-10}$ | NA | NA | -6.40 | $1.5 \times 10^{-10}$ |
| cg07773593 | 19 | 45417793 | ADNI (blood) | *APOC1* | -6.04 | $4.6 \times 10^{-9}$ | -3.00 | $3.6 \times 10^{-3}$ | -4.36 | $2.1 \times 10^{-5}$ |
| cg05644480 | 19 | 45418020 | FHS (blood) | *APOC1* | -5.85 | $5.0 \times 10^{-9}$ | NA | NA | -5.85 | $5.0 \times 10^{-9}$ |

T: T-value, P: P-value. Only CpG sites significantly ( $P < 5 \times 10^{-8}$ ) differentially methylated between *APOE* ε4 carriers and non-carriers in the total sample were included.

**Figure 2.**
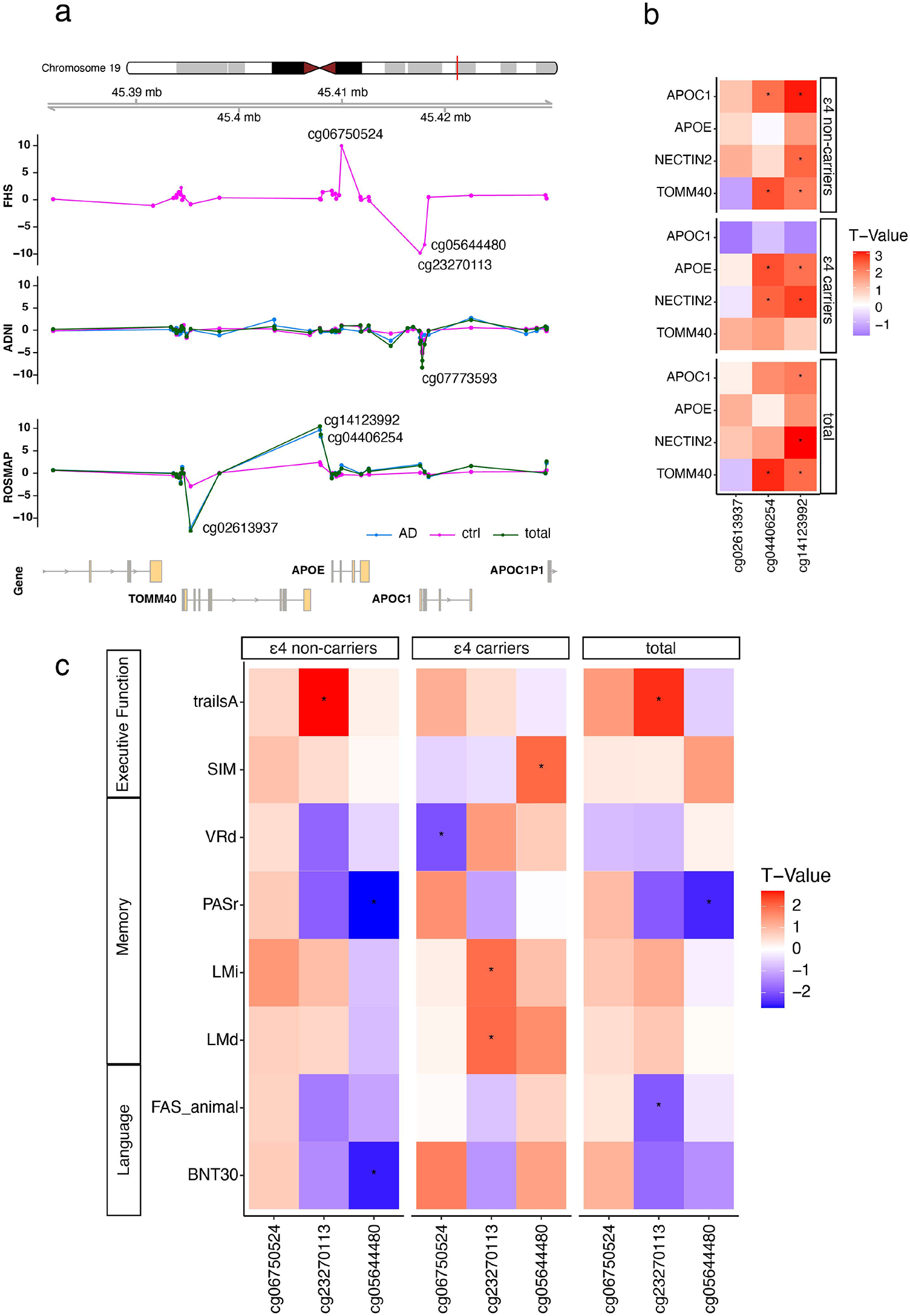
Differential methylation in the *APOE* region between *APOE* ε4 carriers and non-carriers. **(a)** Regional plot of the *APOE* region. Differential methylation between ε4 carriers and non-carriers is shown for the total sample (green line), AD cases (blue line), and controls (pink line) across three datasets. X-axis represents CpG sites that were significantly differentially methylated between *APOE* ε4 carriers and non-carriers at a genome-wide significance level (P<5×10^−8^). Y-axis indicates the log10 P-value of hypermethylation (> 0) or hypomethylation (< 0). **(b)** Heatmap showing association of methylation in brain with expression of *APOE* and adjacent genes. Significant (P<0.05) associations are indicated by an asterisk. **(c)** Heatmap showing association of methylation in blood with cognitive test scores in the FHS dataset. Significant (P<0.05) associations are indicated by an asterisk.

Among the significant CpG sites in the *APOE* region between ε4 carriers and non-carriers in the FHS dataset (**Table 2**), increased methylation in the cg06750524 was associated with poor memory performance measured by the VRd (P=0.04) test in FHS *APOE* ε4 carriers (**Figure 2c and Supplementary Table 5**). Increased methylation of cg23270113 and cg05644480 from the gene *APOC1* was significantly associated with worse performance on trailsA (P=6.9×10^−3^), PASr (P=6.7×10^−3^), and BNT30 (P=0.01) tests in ε4 non-carriers. In *APOE* ε4 carriers, lower methylation at cg23270113 was associated with poor memory performance measured by the LMd (P=0.05) and LMi (P=0.05) tests, and lower methylation at cg05644480 was associated with poor performance on the SIM (P=0.05).

### Co-Methylation Networks

The average methylation level for each of five networks from brain data and three networks from blood data was significantly different between AD and control subjects as well as between ε4 carriers and non-carriers (**Table 3, Supplementary Figure 3, and Supplementary Figure 4**). Five networks (mod2, mod3, mod4, mod5, and mod8) were significantly enriched for eleven pathways (**Figure 3a**). These five networks contained gene-sets including 60 genes that overlap the networks and whose expression levels are modified by estradiol (**Figure 3b and Supplementary Table 6)**. These 60 genes were biologically connected as a subnetwork (**Figure 3c**). Four of these networks (excluding mod5) were enriched for the estrogen response early pathway (**Table 3 and Figure 3a**). *GPR133*, a member of mod2 and mod3 networks, was differentially methylated between AD cases and controls lacking ε4 (P=6.4×10^−6^) and significantly associated with Braak stage (P=5.8×10^−6^) and CERAD score (P=4.6×10^−6^) (**Table 1 and Supplementary Table 2**). Mod5, the only network not enriched for estrogen response early, was uniquely enriched for the E2F target pathway and for seven unique drug perturbation sets (**Table 3, Figure 3a, and Figure 3b**). Mod2 was enriched for estradiol as well as enriched for the complement pathway, mitotic spindles, and TGF-beta signaling (**Figure 3a and Figure 3b**). Mod8 was the only blood network showing significant enrichment for drug perturbations and biological pathways that overlapped significant modules derived from brain (**Figure 3a and Figure 3b**).

**Table 3.** Summary of co-methylated networks from ROSMAP and ADNI associated with AD and *APOE* □4 status

| Module Name | Dataset | Size | AD P-value* | *APOE* P-value* | significant (FDR < 0.05) hallmark pathways | Significant (FDR < 0.05) drug perturbations |
| --- | --- | --- | --- | --- | --- | --- |
| Mod1 | ROSMAP | 288 | 0.03 | $2.0 \times 10^{-4}$ | NA | NA |
| Mod2 | ROSMAP | 6395 | $2.6 \times 10^{-4}$ | $3.1 \times 10^{-5}$ | UV Response Dn, Complement, Epithelial Mesenchymal Transition, Mitotic Spindle, Apical Junction, TGF-beta Signaling, Estrogen Response Early, Coagulation | estradiol |
| Mod3 | ROSMAP | 4891 | $2.9 \times 10^{-3}$ | $6.1 \times 10^{-4}$ | Coagulation, Myogenesis, UV Response Dn, Estrogen Response Early, Epithelial Mesenchymal Transition, Notch Signaling | estradiol, mycophenolate mofetil |
| Mod4 | ROSMAP | 3043 | $2.4 \times 10^{-3}$ | $4.9 \times 10^{-3}$ | Myogenesis, Estrogen Response Early | estradiol, letrozole |
| Mod5 | ROSMAP | 1277 | 0.02 | $1.1 \times 10^{-9}$ | E2F Targets | imatinib, methotrexate, bexarotene, fulvestrant, estradiol, etoposide, tamoxifen, hydroquinone, docetaxel, paclitaxel, valproic acid, ethanol, plicamycin |
| Mod6 | ADNI | 391 | 0.04 | $5.3 \times 10^{-4}$ | NA | NA |
| Mod7 | ADNI | 148 | 0.04 | 0.04 | NA | NA |
| Mod8 | ADNI | 4167 | $4.0 \times 10^{-3}$ | 0.01 | UV Response Dn, Estrogen Response Early, Myogenesis, Apical Junction | doxorubicin, tamoxifen, cisplatin, estradiol, prednisolone, valproic acid, ethanol, rosiglitazone, plicamycin, carboplatin, paclitaxel, hydrocortisone, dexamethasone |

\*AD and *APOE* P-values derived from student's t-test between conditions of module eigenvalues which indicate average methylation over the network.

**Figure 3.**
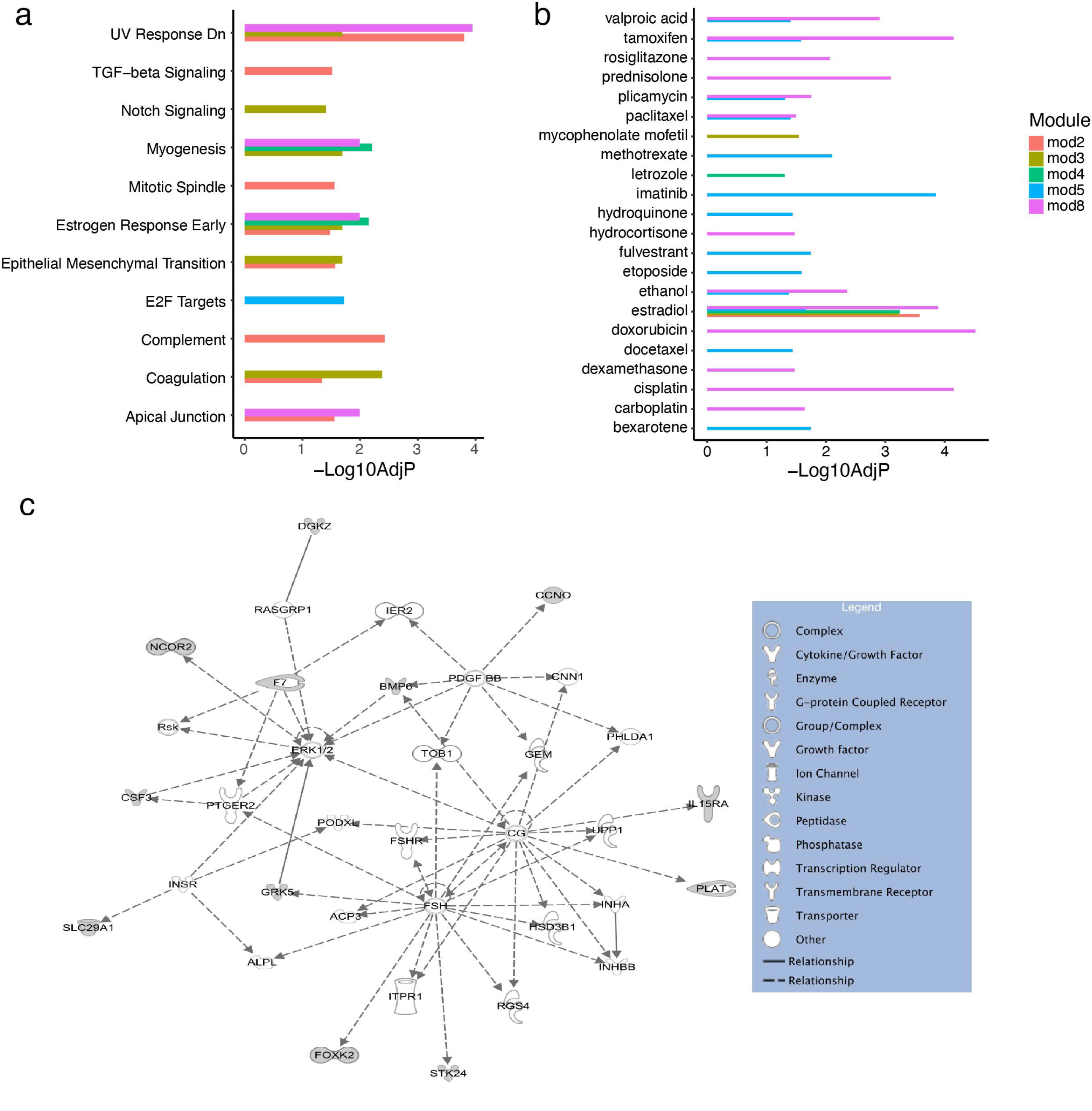
Co-methylation networks. Co-methylation networks included genes with significantly differentially methylated CpG sites (P<0.05) between *APOE* ε4 carriers and non-carriers. Bar plots indicate significant AD and *APOE* genotype-related co-methylation networks containing genes enriched for **(a)** biological pathways and **(b)** drug perturbation gene-sets (i.e., genes whose expression is modified by a drug). Networks with significant pathway or drug gene-set enrichment (adjusted P<0.05) are shown. (c) Biological subnetwork including genes from multiple co-methylation networks enriched for genes whose expression is perturbed by estradiol.

## Discussion

We identified 25 CpG sites in brain and 36 CpG sites in blood that were differentially methylated in AD cases compared to controls in an *APOE* genotype-specific manner. Multiple CpG sites in the *APOE* region were differentially methylated between ε4 carriers and non-carriers in brain or blood. Methylation of several of these CpG sites in blood was significantly associated with performance on cognitive tests in either ε4 carriers or non-carriers. Lastly, we derived eight unique co-methylation networks across blood and brain showing significant differential methylation patterns between AD cases and controls and between ε4 carriers and non-carriers. Five of eight (62.5%) networks included genes enriched for an estradiol drug perturbation gene-set and four of these 5 networks were involved in estrogen response pathway. These findings suggest that AD-related methylation patterns are dependent on *APOE* genotype and may be targeted by estrogen modulating drugs.

CpG sites in *GPR133* and *CHSY1* were hypermethylated in brain from AD cases lacking *APOE* ε4 and associated with measures of plaque and tangle pathology. *GPR133* is a member of the adhesion G protein-coupled receptor family, several of which have been implicated in AD and proposed as potential drug targets for neurological disease [26]. A deletion in *CHSY1* causes an increased inflammatory response and hippocampal neurodegeneration in mice [27]. We also observed blood hypomethylation at CpG sites from the *CHEK1* gene in AD cases carrying *APOE* ε4. *CHEK1* induces astrogliosis in AD brains and inhibits PP2A which was linked in *APOE* genotype-specific patterns to AD and AD-related traits by GWAS, gene expression analysis, and experimental studies [6,28].

Association of AD with variants in the *APOE* region has been extensively evaluated [29]. While the link between *APOE* isoforms and AD risk is well established, independent associations for AD with other genes near the *APOE* gene including *TOMM40* and *APOC1* are less conclusive because they often do not replicate across ethnic populations and are confounded by high linkage disequilibrium with *APOE* variants [30,31]. However, methylation studies have consistently shown unique and strong differential methylation patterns by *APOE* genotype in AD cases and controls [11,12]. We confirmed decreased methylation on *APOC1* in blood and increased methylation on *APOE* in brain among ε4 carriers compared to non-carriers, while decreased methylation of ε4 carriers on *TOMM40* in brain. These results suggest possible distinct contributions of the genes in the *APOE* region between blood and brain tissues through differential regulation on methylation sites to AD risk. Future studies are necessary to understand the exact mechanisms involved with methylation and AD between blood and brain tissues in an *APOE* genotype-specific manner.

We identified pathways enriched for genes in *APOE* genotype and AD-specific co-methylation networks that were derived from differentially methylated CpG sites between ε4 carriers and non-carriers in brain or blood. One of the brain networks was enriched for genes in complement pathway that was previously linked to AD in an *APOE* genotype-specific manner [5,6]. Five of the eight *APOE* ε4 associated networks showed significant enrichment with genes perturbed by a drug, estradiol. Estradiol has been associated with increased cognitive function in both animals and humans [32]. Loss of estrogen in post-menopausal women has been associated with increased AD risk [33] and estrogen replacement therapy has shown to decrease AD risk in post-menopausal women [34], particularly among those under age 64 [35]. Furthermore, the effect of estrogen use on AD risk may be limited to ε4 non-carriers [36]. A recent study showed estrogen decreased amyloid-β accumulation in the hippocampus and cortex in mice lacking ε4 [37].

Our study has several limitations. First, methylation data from a microarray platform in ADNI participants was different from the one used by FHS and ROSMAP. Therefore, direct replication of each CpG site among these studies was limited. Second, none of the FHS participants with methylation data had AD due to their relatively young age. Alternatively, we evaluated cognitive performance data which served as a robust AD endophenotype. Third, pathway enrichment analysis was conducted using genes with network CpG sites under the assumption of each CpG site directly modulating the corresponding gene. This assumption may not hold when the CpG site regulates a long-distant gene. Fourth, phenotype data were not comparable across datasets; in particular, neuropathological measures (i.e., Braak stage and CERAD score) were available for ROSMAP, whereas ADNI and FHS featured cognitive test data. Finally, the datasets included in this study were too small to account for sex differences after stratification by *APOE* genotype or AD status. As a result, we were unable to evaluate sex effect, especially genes involved in estrogen response pathways.

In conclusion, we identified differentially methylated CpG sites in many genes including *APOE* that were also associated with AD and related traits. Many of these associations were *APOE* genotype- or tissue-specific. AD and *APOE* genotype-specific methylation networks were linked to estrogen response and an estrogen replacement therapy, estradiol. Future studies are required to evaluate the contributions of methylation and *APOE* genotypes to beneficial effects of estrogen as an AD risk-lowering therapy.

## Data Availability

FHS data are available on the dbGaP (Study Accession ID:  phs000056.v5.p3). ROSMAP resources can be requested at from the CommonMind Consortium portal (http://www.synapse.org). Data used in preparation of this article were obtained from the Alzheimer Disease Neuroimaging Initiative (ADNI) database (http://adni.loni.usc.edu). As such, the investigators within the ADNI contributed to the design and implementation of ADNI and/or provided data but did not participate in analysis or writing of this report. A complete listing of ADNI investigators can be found at: http://adni.loni.usc.edu/wp-content/uploads/how_to_apply/ADNI_Acknowledgement_List.pdf.

http://adni.loni.usc.edu

## Abbreviations

AD: Alzheimer’s Disease
ADAS13: Alzheimer’s Disease Assessment Scale – 13 item
ADNI: Alzheimer’s Disease Neuroimaging Initiative
BNT30: Boston Naming Test
CDRSB: Clinical Dementia Rating Scale Sum of Boxes
CERAD: Consortium to Establish a Registry for Alzheimer Disease
CpG: cytosine-phosphate-guanine
FAS_animal: animal portion of the Verbal Fluency Test
FHS: Framingham Heart Study
GRM: genetic relatedness matrix
GWAS: genome-wide association study
LDELTOTAL: logical memory - delayed recall
LMd: Logical Memories – Delayed Recall
LMi: Logical Memories – Immediate Recall
MCI: mild cognitive impairment
MRI: magnetic resonance imaging
PASr: Paired Associate Learning – Recognition
PMI: post mortem interval
RAVLT: Rey Auditory Verbal Learning Test
RIN: RNA integrity number
ROSMAP: Religious Orders Study and Rush Memory and Aging Project
RNA-seq: RNA sequencing
SIM: Similarities Test
trailsA: Trail A
VRd: Visual Reproductions – Delayed Recall
WGCNA: weighted gene correlation network analysis

## Acknowledgements

Not applicable

## Availability of data and materials

FHS data are available on the dbGaP (Study Accession ID: phs000056.v5.p3). ROSMAP resources can be requested at from the CommonMind Consortium portal (http://www.synapse.org). Data used in preparation of this article were obtained from the Alzheimer’s Disease Neuroimaging Initiative (ADNI) database (http://adni.loni.usc.edu). As such, the investigators within the ADNI contributed to the design and implementation of ADNI and/or provided data but did not participate in analysis or writing of this report. A complete listing of ADNI investigators can be found at: http://adni.loni.usc.edu/wp-content/uploads/how_to_apply/ADNI_Acknowledgement_List.pdf.

## Authors’ contributions

R.P., L.A.F., and G.R.J. conceived overall study design. R.P., K.L., and G.R.J. perceived statistical analysis. R.P., N.P., and J.H. performed data analyses. D.A.B., K.N., and R.A. provided cognitive test and imaging data as well as interpretation of the data from the ROSMAP, ADNI, and FHS, respectively. T.D.S. conducted and provided neuropathological data using autopsied brains. R.P., K.N., D.A.B., T.D.S., L.A.F., and G.R.J. reviewed and edited the manuscript. G.R.J. and L.A.F. supervised and obtained funding for the project.

## Ethics approval and consent to participate

The study protocol, design, and performance of the current study were approved by the Boston University Institutional Review Board.

## Consent for publication

Not applicable

## References

1. DeTure MA, Dickson DW. The neuropathological diagnosis of Alzheimer’s disease. Molecular Neurodegeneration. 2019;14:32.

2. Farrer LA, Cupples LA, Haines JL, Hyman B, Kukull WA, Mayeux R, et al. Effects of age, sex, and ethnicity on the association between apolipoprotein E genotype and Alzheimer disease. A meta-analysis. APOE and Alzheimer Disease Meta Analysis Consortium. JAMA. 1997;278:1349–1356.

3. Corder EH, Saunders AM, Strittmatter WJ, Schmechel DE, Gaskell PC, Small GW, et al. Gene dose of apolipoprotein E type 4 allele and the risk of Alzheimer’s disease in late onset families. Science. 1993;261:921–923.

4. Reiman EM, Arboleda-Velasquez JF, Quiroz YT, Huentelman MJ, Beach TG, Caselli RJ, et al. Exceptionally low likelihood of Alzheimer’s dementia in APOE2 homozygotes from a 5,000-person neuropathological study. Nat Commun. 2020;11:1–11.

5. Panitch R, Hu J, Chung J, Zhu C, Meng G, Xia W, et al. Integrative brain transcriptome analysis links complement component 4 and HSPA2 to the APOE ε2 protective effect in Alzheimer disease. Mol Psychiatry. 2021:1–11.

6. Jun GR, You Y, Zhu C, Meng G, Chung J, Panitch R, et al. Protein phosphatase 2A and complement component 4 are linked to the protective effect of APOE LJ2 for Alzheimer’s disease. Alzheimers Dement. 2022. 9 February 2022. https://doi.org/10.1002/alz.12607.

7. Panitch R, Hu J, Xia W, Bennett DA, Stein TD, Farrer LA, et al. Blood and brain transcriptome analysis reveals APOE genotype-mediated and immune-related pathways involved in Alzheimer disease. Alzheimers Res Ther. 2022;14:30.

8. Bellenguez C, Küçükali F, Jansen IE, Kleineidam L, Moreno-Grau S, Amin N, et al. New insights into the genetic etiology of Alzheimer’s disease and related dementias. Nat Genet. 2022;54:412–436.

9. De Jager P, Srivastava G, Lunnon K, Burgess J, Schalkwyk L, Yu L, et al. Alzheimery’s disease pathology is associated with early alterations in brain DNA methylation at ANK1, BIN1, RHBDF2 and other loci. Nat Neurosci. 2014;17:1156–1163.

10. Li QS, Vasanthakumar A, Davis JW, Idler KB, Nho K, Waring JF, et al. Association of peripheral blood DNA methylation level with Alzheimer’s disease progression. Clinical Epigenetics. 2021;13:191.

11. Foraker J, Millard SP, Leong L, Thomson Z, Chen S, Keene CD, et al. The APOE Gene is Differentially Methylated in Alzheimer’s Disease. J Alzheimers Dis. 2015;48:745–755.

12. Walker RM, Vaher K, Bermingham ML, Morris SW, Bretherick AD, Zeng Y, et al. Identification of epigenome-wide DNA methylation differences between carriers of APOE ε4 and APOE ε2 alleles. Genome Medicine. 2021;13:1.

13. Yu L, Lutz MW, Wilson RS, Burns DK, Roses AD, Saunders AM, et al. TOMM40′523 variant and cognitive decline in older persons with APOE ε3/3 genotype. Neurology. 2017;88:661–668.

14. De Jager PL, Ma Y, McCabe C, Xu J, Vardarajan BN, Felsky D, et al. A multi-omic atlas of the human frontal cortex for aging and Alzheimer’s disease research. Sci Data. 2018;5:180142.

15. Bennett DA, Buchman AS, Boyle PA, Barnes LL, Wilson RS, Schneider JA. Religious Orders Study and Rush Memory and Aging Project. J Alzheimers Dis. 2018;64:S161–S189.

16. Hyman BT, Phelps CH, Beach TG, Bigio EH, Cairns NJ, Carrillo MC, et al. National Institute on Aging–Alzheimer’s Association guidelines for the neuropathologic assessment of Alzheimer’s disease. Alzheimers Dement. 2012;8:1–13.

17. Braak H, Braak E. Neuropathological stageing of Alzheimer-related changes. Acta Neuropathol. 1991;82:239–259.

18. Mirra SS, Heyman A, McKeel D, Sumi SM, Crain BJ, Brownlee LM, et al. The Consortium to Establish a Registry for Alzheimer’s Disease (CERAD). Part II. Standardization of the neuropathologic assessment of Alzheimer’s disease. Neurology. 1991;41:479–486.

19. Pidsley R, Y Wong Cc, Volta M, Lunnon K, Mill J, Schalkwyk LC. A data-driven approach to preprocessing Illumina 450K methylation array data. BMC Genomics. 2013;14:293.

20. Splansky GL, Corey D, Yang Q, Atwood LD, Cupples LA, Benjamin EJ, et al. The Third Generation Cohort of the National Heart, Lung, and Blood Institute’s Framingham Heart Study: Design, Recruitment, and Initial Examination. American Journal of Epidemiology. 2007;165:1328–1335.

21. Ritchie ME, Phipson B, Wu D, Hu Y, Law CW, Shi W, et al. limma powers differential expression analyses for RNA-sequencing and microarray studies. Nucleic Acids Res. 2015;43:e47–e47.

22. Yang J, Lee SH, Goddard ME, Visscher PM. GCTA: a tool for genome-wide complex trait analysis. Am J Hum Genet. 2011;88:76–82.

23. Jun G, Guo H, Klein BEK, Klein R, Wang JJ, Mitchell P, et al. EPHA2 Is Associated with Age-Related Cortical Cataract in Mice and Humans. PLoS Genet. 2009;5:e1000584.

24. Langfelder P, Horvath S. WGCNA: an R package for weighted correlation network analysis. BMC Bioinformatics. 2008;9:559.

25. Chen EY, Tan CM, Kou Y, Duan Q, Wang Z, Meirelles GV, et al. Enrichr: interactive and collaborative HTML5 gene list enrichment analysis tool. BMC Bioinformatics. 2013;14:128.

26. Folts CJ, Giera S, Li T, Piao X. Adhesion G protein-coupled receptors as drug target for neurological diseases. Trends Pharmacol Sci. 2019;40:278–293.

27. Macke EL, Henningsen E, Jessen E, Zumwalde NA, Landowski M, Western DE, et al. Loss of Chondroitin Sulfate Modification Causes Inflammation and Neurodegeneration in skt Mice. Genetics. 2020;214:121–134.

28. Zhou Y, Liu X, Ma S, Zhang N, Yang D, Wang L, et al. ChK1 activation induces reactive astrogliosis through CIP2A/PP2A/STAT3 pathway in Alzheimer’s disease. FASEB J. 2022;36:e22209.

29. Jun G, Vardarajan BN, Buros J, Yu C-E, Hawk MV, Dombroski BA, et al. Comprehensive Search for Alzheimer Disease Susceptibility Loci in the APOE Region. Arch Neurol. 2012;69:1270–1279.

30. Zhou Q, Zhao F, Lv Z, Zheng C, Zheng W, Sun L, et al. Association between APOC1 Polymorphism and Alzheimer’s Disease: A Case-Control Study and Meta-Analysis. PLoS One. 2014;9:e87017.

31. Chiba-Falek O, Gottschalk WK, Lutz MW. The effects of the TOMM40 poly-T alleles on Alzheimer’s disease phenotypes. Alzheimers Dement. 2018;14:692–698.

32. Luine VN. Estradiol and cognitive function: Past, present and future. Horm Behav. 2014;66:602–618.

33. Ratnakumar A, Zimmerman SE, Jordan BA, Mar JC. Estrogen activates Alzheimer’s disease genes. Alzheimers Dement (N Y). 2019;5:906–917.

34. Song Y, Li S, Li X, Chen X, Wei Z, Liu Q, et al. The Effect of Estrogen Replacement Therapy on Alzheimer’s Disease and Parkinson’s Disease in Postmenopausal Women: A Meta-Analysis. Frontiers in Neuroscience. 2020;14.

35. Espeland MA, Shumaker SA, Leng I, Manson JE, Brown CM, LeBlanc ES, et al. Long Term Effects on Cognitive Function of Postmenopausal Hormone Therapy Prescribed to Women Aged 50–55 Years. JAMA Intern Med. 2013;173:10.1001/jamainternmed.2013.7727.

36. Yaffe K, Haan M, Byers A, Tangen C, Kuller L. Estrogen use, APOE, and cognitive decline: evidence of gene-environment interaction. Neurology. 2000;54:1949–1954.

37. Kunzler J, Youmans KL, Yu C, LaDu MJ, Tai L. APOE modulates the effect of estrogen therapy on Aβ accumulation EFAD-Tg mice. Neurosci Lett. 2014;560:131–136.

