## Supplementary Materials for "*APOE* Genotype-specific Methylation Patterns are Linked to Alzheimer Disease Pathology and Estrogen Response"

### SUPPLEMENTARY INFORMATION

**Supplementary Table 1.** Methylation Array Sample Information

| Dataset | APOE Genotype | AD Cases | | | Controls | | |
|---|---|---|---|---|---|---|---|
| | | N | % Female | Age | N | % Female | Age |
| ROSMAP | ε4 non-carriers | 265 | 69.4 | 90.2 ± 5.7 | 246 | 58.9 | 86.4 ± 7.3 |
| | ε4 carriers | 152 | 62.5 | 87.8 ± 5.8 | 34 | 55.9 | 84.6 ± 6.7 |
| | TOTAL | 417 | 66.8 | 89.3 ± 5.9 | 280 | 58.6 | 86.1 ± 7.2 |
| ADNI | ε4 non-carriers | 29 | 34.5 | 78.3 ± 7.1 | 158 | 50.0 | 74.9 ± 5.9 |
| | ε4 carriers | 62 | 35.5 | 72.9 ± 7.0 | 52 | 50.0 | 74.1± 5.4 |
| | TOTAL | 91 | 35.2 | 74.6 ± 7.4 | 210 | 50.0 | 74.7 ± 5.8 |
| FHS | ε4 non-carriers | NA | NA | NA | 1115 | 45.9 | 45.9 ± 8.2 |
| | ε4 carriers | NA | NA | NA | 276 | 50.4 | 45.7 ± 8.5 |
| | TOTAL | NA | NA | NA | 1391 | 52.3 | 45.8 ± 8.2 |

ROSMAP: Religious Orders Study and Rush Memory and Aging Project; ADNI: Alzheimer's Disease Neuroimaging Initiative; FHS: Framingham Heart Study. Methylation array data were generated from autopsied brains in the ROSMAP and blood in the ADNI and FHS.

**Supplementary Table 2.** Association of AD differentially methylated CpG Sites ($P<10^{-5}$) from *APOE* ε4 carriers and non-carriers in brain with tangle and plaque pathology

| CpG Name | Chr | Position | Gene | Braak Stage | | CERAD Score | |
|---|---|---|---|---|---|---|---|
| | | | | T | P | T | P |
| cg19533050 | 2 | 163175044 | IFIH1 | -1.26 | 0.21 | 0.22 | 0.82 |
| cg23808213 | 2 | 166948291 | SCN1A | -2.74 | $6.3x10^{-3}$ | 3.73 | $2.1x10^{-4}$ |
| cg05731218 | 2 | 216769199 | | -6.99 | $6.6x10^{-12}$ | 6.10 | $1.8x10^{-9}$ |
| cg04436449 | 3 | 185214835 | TMEM41A | -2.66 | $7.9x10^{-3}$ | 3.42 | $6.6x10^{-4}$ |
| cg12307200 | 3 | 188664632 | | -7.05 | $4.2x10^{-12}$ | 7.36 | $5.1x10^{-13}$ |
| cg16234490 | 4 | 77138082 | FAM47E | -2.78 | $5.6x10^{-3}$ | 3.54 | $4.3x10^{-4}$ |
| cg24899806 | 7 | 119914282 | KCND2 | -2.15 | 0.03 | 3.28 | $1.1x10^{-3}$ |
| cg23831517 | 8 | 34182528 | | 2.69 | $7.4x10^{-3}$ | -3.51 | $4.7x10^{-4}$ |
| cg14096074 | 9 | 34255149 | KIF24 | -2.14 | 0.03 | 3.34 | $8.7x10^{-4}$ |
| cg03727169 | 10 | 31418969 | | -2.81 | $5.1x10^{-3}$ | 2.54 | 0.01 |
| cg01982597 | 10 | 50733420 | ERCC6 | -3.43 | $6.3x10^{-4}$ | 3.26 | $1.1x10^{-3}$ |
| cg20326704 | 10 | 70321770 | TET1 | -1.86 | 0.06 | 0.86 | 0.39 |
| cg04126866 | 10 | 85932763 | C10orf99 | -3.89 | $1.1x10^{-4}$ | 4.43 | $1.1x10^{-5}$ |
| cg14882481 | 11 | 107437051 | ALKBH8 | -2.16 | 0.03 | 4.02 | $6.4x10^{-5}$ |
| cg10907744 | 12 | 131589455 | GPR133 | 4.57 | $5.8x10^{-6}$ | -4.62 | $4.6x10^{-6}$ |
| cg18708502 | 13 | 21588555 | LATS2 | 2.95 | $3.3x10^{-3}$ | -3.38 | $7.7x10^{-4}$ |
| cg16746221 | 14 | 20666088 | OR11G2 | -2.59 | 0.01 | 3.03 | $2.5x10^{-3}$ |
| cg24231804 | 15 | 67316861 | | -3.07 | $2.2x10^{-3}$ | 4.27 | $2.2x10^{-5}$ |
| cg14829066 | 15 | 88559141 | NTRK3 | 3.33 | $9.2x10^{-4}$ | -4.41 | $1.2x10^{-5}$ |
| cg19987111 | 15 | 101747167 | CHSY1 | -5.41 | $8.5x10^{-8}$ | 4.39 | $1.3x10^{-5}$ |
| cg02432274 | 16 | 88378468 | | 2.53 | 0.01 | -1.45 | 0.15 |
| cg05952786 | 17 | 48559485 | RSAD1 | -2.42 | 0.02 | 3.96 | $8.4x10^{-5}$ |
| cg15503752 | 17 | 74639731 | ST6GALNAC1 | 3.94 | $9.1x10^{-5}$ | -3.56 | $4.0x10^{-4}$ |
| cg05421550 | 19 | 4446485 | UBXN6 | -1.53 | 0.13 | 3.89 | $1.1x10^{-4}$ |
| cg19612770 | 19 | 4475216 | HDGF2 | -2.74 | $6.4x10^{-3}$ | 3.87 | $1.2x10^{-4}$ |

T: T-value, P: P-value

**Supplementary Table 3.** AD differentially methylated CpG Sites ($P<10^{-5}$) from *APOE* ε4 carriers and non-carriers in blood

| CpG Name | Chr | Position | Gene | Total Sample | | *APOE* ε4 carriers | | *APOE* ε4 non-carriers | |
|---|---|---|---|---|---|---|---|---|---|
| | | | | T | P | T | P | T | P |
| cg09825488 | 1 | 40974006 | EXO5 | 3.95 | $9.7x10^{-5}$ | 0.68 | 0.50 | 4.84 | $2.9x10^{-6}$ |
| cg21836919 | 1 | 207351463 | | -2.85 | $4.6x10^{-3}$ | -4.68 | $8.6x10^{-6}$ | 1.00 | 0.32 |
| cg25631371 | 1 | 229846417 | | -2.80 | $5.5x10^{-3}$ | -0.58 | 0.56 | -4.80 | $3.3x10^{-6}$ |
| cg21055045 | 2 | 47266465 | TTC7A | 4.20 | $3.5x10^{-5}$ | 5.02 | $2.1x10^{-6}$ | 0.87 | 0.38 |
| cg01447263 | 2 | 54366321 | ACYP2 | 2.96 | $3.3x10^{-3}$ | 4.78 | $5.8x10^{-6}$ | 0.22 | 0.82 |
| cg15739581 | 2 | 166626783 | GALNT3 | -3.34 | $9.4x10^{-4}$ | -4.79 | $5.5x10^{-6}$ | 0.52 | 0.60 |
| cg05303734 | 2 | 178101031 | NFE2L2 | -2.61 | $9.4x10^{-3}$ | -0.16 | 0.88 | -4.77 | $3.9x10^{-6}$ |
| cg00361562 | 2 | 198649771 | BOLL | -1.99 | 0.05 | 0.85 | 0.40 | -4.66 | $6.2x10^{-6}$ |
| cg19577697 | 2 | 232123245 | ARMC9 | 2.33 | 0.02 | -0.72 | 0.47 | 4.56 | $9.5x10^{-6}$ |
| cg10420726 | 4 | 71599662 | RUFY3 | 1.93 | 0.05 | 4.70 | $8.0x10^{-6}$ | -0.73 | 0.47 |
| cg05845376 | 5 | 140683632 | SLC25A2 | -2.77 | $5.9x10^{-3}$ | -5.04 | $2.0x10^{-6}$ | 1.05 | 0.29 |
| cg15595495 | 6 | 29798726 | HLA-G | -1.36 | 0.17 | -4.74 | $6.6x10^{-6}$ | 0.89 | 0.38 |
| cg00198525 | 6 | 74165863 | | -2.30 | 0.02 | -4.90 | $3.5x10^{-6}$ | 0.72 | 0.47 |
| cg06002867 | 6 | 170449679 | | -1.86 | 0.06 | 0.92 | 0.36 | -5.11 | $8.3x10^{-7}$ |
| cg09236008 | 7 | 23246922 | | -2.36 | 0.02 | -4.84 | $4.4x10^{-6}$ | -0.29 | 0.77 |
| cg17865549 | 8 | 23541133 | NKX3-1 | -2.41 | 0.02 | -4.64 | $1.0x10^{-5}$ | 0.40 | 0.69 |
| cg22524508 | 11 | 13461822 | BTBD10 | -1.29 | 0.20 | 0.95 | 0.34 | -4.60 | $8.0x10^{-6}$ |
| cg10370850 | 11 | 70027093 | ANO1 | 3.33 | $9.7x10^{-4}$ | 4.88 | $3.8x10^{-6}$ | 1.23 | 0.22 |
| cg18714484 | 11 | 125511817 | CHEK1 | -4.09 | $5.5x10^{-5}$ | -5.01 | $2.2x10^{-6}$ | -0.38 | 0.71 |
| cg26362686 | 12 | 103125021 | | -2.39 | 0.02 | -0.21 | 0.83 | -5.38 | $2.3x10^{-7}$ |
| cg25190513 | 12 | 123201362 | GPR109B | -3.09 | $2.2x10^{-3}$ | -4.68 | $8.6x10^{-6}$ | -0.22 | 0.82 |
| cg24154450 | 14 | 28851873 | | 2.37 | 0.02 | 5.29 | $6.8x10^{-7}$ | -0.42 | 0.68 |
| cg05890727 | 14 | 55764647 | FBXO34 | -2.76 | $6.1x10^{-3}$ | -4.81 | $5.0x10^{-6}$ | 0.41 | 0.68 |
| cg14297991 | 14 | 97014011 | PAPOLA | 1.39 | 0.17 | 5.18 | $1.1x10^{-6}$ | -2.02 | 0.05 |
| cg07372700 | 15 | 31281399 | MTMR10 | 2.99 | $3.0x10^{-3}$ | 4.78 | $5.7x10^{-6}$ | -1.04 | 0.30 |
| cg21058182 | 15 | 90401931 | AP3S2 | -1.99 | 0.05 | 0.74 | 0.46 | -4.56 | $9.5x10^{-6}$ |
| cg14381313 | 16 | 88268339 | | -3.68 | $2.8x10^{-4}$ | -0.12 | 0.91 | -5.65 | $6.4x10^{-8}$ |
| cg08951186 | 16 | 88290370 | | -2.82 | $5.2x10^{-3}$ | -0.47 | 0.64 | -4.56 | $9.4x10^{-6}$ |
| cg13785068 | 17 | 3416273 | | -2.90 | $4.0x10^{-3}$ | -0.32 | 0.75 | -4.57 | $9.3x10^{-6}$ |
| cg18493677 | 17 | 3416525 | TRPV3 | -2.63 | $8.9x10^{-3}$ | 0.04 | 0.97 | -4.88 | $2.3x10^{-6}$ |
| cg23018236 | 17 | 30244563 | | -2.73 | $6.8x10^{-3}$ | -5.20 | $9.8x10^{-7}$ | 0.70 | 0.49 |
| cg12989128 | 20 | 42875933 | GDAP1L1 | 2.39 | 0.02 | -0.52 | 0.60 | 4.75 | $4.2x10^{-6}$ |
| cg22522263 | 21 | 23391891 | LOC1019 27843 | 1.19 | 0.23 | 4.92 | $3.3x10^{-6}$ | -1.97 | 0.05 |
| cg01196038 | 21 | 37985032 | | -2.98 | $3.2x10^{-3}$ | 0.53 | 0.60 | -4.67 | $5.8x10^{-6}$ |
| cg03172077 | 22 | 22402314 | | -2.87 | $4.4x10^{-3}$ | -4.91 | $3.4x10^{-6}$ | 1.05 | 0.29 |
| cg23304135 | X | 3187835 | | 1.65 | 0.10 | 4.67 | $8.9x10^{-6}$ | -1.04 | 0.30 |

Differential methylation was carried out in the total sample, *APOE* ε4 carriers, and non-carriers
T: T-value, P: P-value

**Supplementary Table 4.** Association of AD differentially methylated CpG Sites ($P<10^{-5}$) from *APOE* ε4 carriers and non-carriers in blood with brain imaging and cognitive traits

| CpG Name | Chr | Position | Gene | Most Significant Global Cognitive | | Most Significant Domain-Specific Cognitive | | Most Significant Imaging | |
|---|---|---|---|---|---|---|---|---|---|
| | | | | Test | P | Test | P | Test | P |
| cg09825488 | 1 | 40974006 | EXO5 | CDRSB | $3.3x10^{-6}$ | RAVLT_immediate | $1.9x10^{-4}$ | Hippocampal Volume | $5.6x10^{-3}$ |
| cg21836919 | 1 | 207351463 | | ADAS13 | 0.01 | RAVLT_perc_forgetting | $2.8x10^{-4}$ | Entorhinal Thickness | 0.57 |
| cg21055045 | 2 | 47266465 | TTC7A | ADAS13 | $9.7x10^{-4}$ | RAVLT_perc_forgetting | $3.4x10^{-4}$ | Hippocampal Volume | $4.2x10^{-3}$ |
| cg15739581 | 2 | 166626783 | GALNT3 | ADAS13 | $1.3x10^{-3}$ | RAVLT_perc_forgetting | $4.1x10^{-5}$ | Entorhinal Thickness | 0.03 |
| cg00198525 | 6 | 74165863 | | ADAS13 | $9.0x10^{-3}$ | RAVLT_perc_forgetting | $1.3x10^{-3}$ | Hippocampal Volume | $1.5x10^{-4}$ |
| cg17865549 | 8 | 23541133 | NKX3-1 | ADAS13 | 0.05 | RAVLT_perc_forgetting | $6.8x10^{-4}$ | Ventricle Volume | 0.06 |
| cg10370850 | 11 | 70027093 | ANO1 | ADAS13 | $3.9x10^{-4}$ | RAVLT_learning | $2.8x10^{-3}$ | Ventricle Volume | 0.06 |
| cg18714484 | 11 | 125511817 | CHEK1 | ADAS13 | $1.8x10^{-5}$ | RAVLT_perc_forgetting | $4.2x10^{-5}$ | Ventricle Volume | $8.1x10^{-4}$ |
| cg25190513 | 12 | 123201362 | GPR109B | ADAS13 | $7.4x10^{-4}$ | RAVLT_perc_forgetting | $1.3x10^{-4}$ | Entorhinal Thickness | $3.4x10^{-4}$ |
| cg05890727 | 14 | 55764647 | FBXO34 | ADAS13 | 0.08 | RAVLT_perc_forgetting | $3.8x10^{-5}$ | Entorhinal Thickness | 0.14 |
| cg14381313 | 16 | 88268339 | | ADAS13 | $9.0x10^{-5}$ | RAVLT_perc_forgetting | $1.3x10^{-4}$ | Hippocampal Volume | $1.5x10^{-3}$ |
| cg13785068 | 17 | 3416273 | | CDRSB | $1.4x10^{-3}$ | LDELTOTAL | $2.1x10^{-3}$ | Hippocampal Volume | 0.05 |
| cg23018236 | 17 | 30244563 | | ADAS13 | $8.7x10^{-4}$ | RAVLT_immediate | $2.0x10^{-3}$ | Entorhinal Thickness | 0.01 |
| cg01196038 | 21 | 37985032 | | ADAS13 | $1.1x10^{-3}$ | RAVLT_immediate | $8.0x10^{-4}$ | Ventricle Volume | 0.13 |
| cg03172077 | 22 | 22402314 | | ADAS13 | 0.02 | RAVLT_perc_forgetting | $4.5x10^{-4}$ | Entorhinal Thickness | $2.3x10^{-3}$ |

Only CpG islands with methylation significantly ($p<1.4x10^{-3}$) association with at least one trait shown

**Supplementary Table 5.** Association of significant differentially *APOE* methylated CpGs from the APOE region in FHS with cognitive traits

| Study and CpG Name | Chr | Position | Gene | Most Significant Cognitive Test | P |
|---|---|---|---|---|---|
| **Total sample:** | | | | | |
| cg06750524 | 19 | 45409955 | APOE | trailsA | 0.16 |
| cg23270113 | 19 | 45417587 | APOC1 | trailsA | 0.01 |
| cg05644480 | 19 | 45418020 | APOC1 | PASr | 0.01 |
| **APOE e4 carriers:** | | | | | |
| cg06750524 | 19 | 45409955 | APOE | VRd | 0.04 |
| cg23270113 | 19 | 45417587 | APOC1 | LMd | 0.05 |
| cg05644480 | 19 | 45418020 | APOC1 | SIM | 0.04 |
| **APOE e4 non-carriers:** | | | | | |
| cg06750524 | 19 | 45409955 | APOE | LMi | 0.15 |
| cg23270113 | 19 | 45417587 | APOC1 | trailsA | $6.9x10^{-3}$ |
| cg05644480 | 19 | 45418020 | APOC1 | PASr | $6.7x10^{-3}$ |

**Supplementary Table 6. Overlapping Genes from significant co-methylated networks with Estradiol perturbation**

| Module Name | Dataset | Genes |
|---|---|---|
| Mod2 | ROSMAP | CSF3;PLAT;KLHDC7B; CDH4;HOXA3; EPHB1; SOX5; IL15RA;TLE2;DAPK2; KRT9; CCNO; EMID2;SHC2;MFI2; LTBP3; CHST11; MBP; KCNN4; EXPH5;RAP1GAP2; PTPRN2;BTBD11; BMP6; LYPD6B; DOCK9; PIGZ; MYOM2;PITPNC1; TRIM7;NHSL1; GPR133; CAMK1D;HEG1;NAV1;MANEAL;F7; KCNMA1;GPR126;SHANK2; TNXB; PDE11A; FLNB; OPCML; C1ORF21;C15ORF27;HLA-DPB2 |
| Mod3 | ROSMAP | CSF3;PLAT;KLHDC7B;CDH4; HOXA3; EPHB1;SOX5;IL15RA;TLE2; DAPK2;KRT9;CCNO;EMID2;SHC2; MFI2;LTBP3; MBP; KCNN4;EXPH5;RAP1GAP2;PTPRN2; SLC4A11;BTBD11;BMP6;LYPD6B;CRYL1; DOCK9;PIGZ; MYOM2; TRIM7;NHSL1; GPR133; CAMK1D;HEG1; NAV1;MANEAL;F7; KCNMA1;GPR126; SHANK2;TNXB; PDE11A; FLNB;OPCML;C15ORF27; HLA-DPB2 |
| Mod4 | ROSMAP | SLC16A3;DGKZ;GRK5; STK24; VWF; NCOR2; SLC29A1; TNXB; FOXK2; PPM1F; FLNB; CYB561 |
| Mod5 | ROSMAP | NCOA4 |
| Mod8 | ADNI | PLAT;SLC16A3;DAPK2;DGKZ;GRK5;CHST11;STK24;PITPNC1;VWF;NCOA4; NCOR2;SLC29A1;SHANK2; FOXK2;PPM1F; FLNB;C1ORF21;CYB561 |

**Supplementary Figure 1: AD differentially methylated CpGs in Blood in Brain by *APOE* genotype**

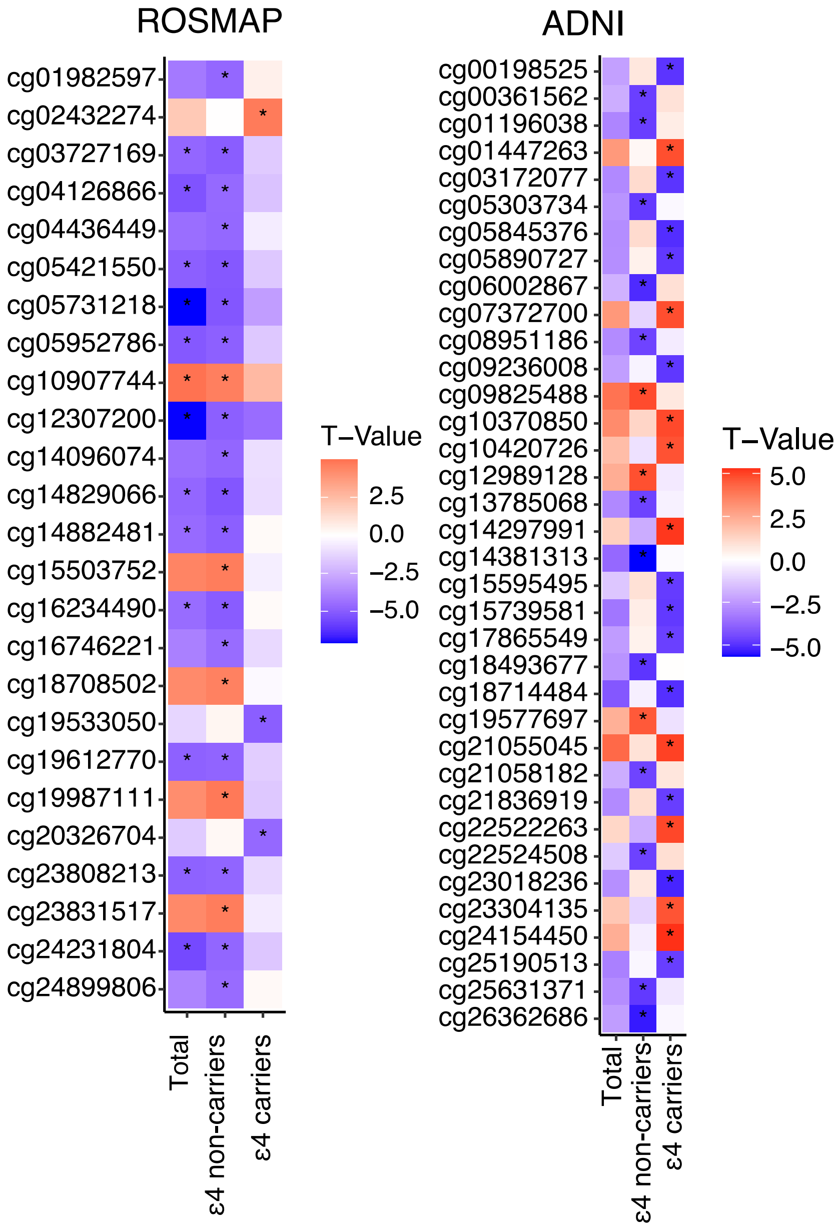

**Supplementary Figure 2: Heatmap associations with nearby gene expression for significant differentially CpGs in ADNI (cg07773593)**

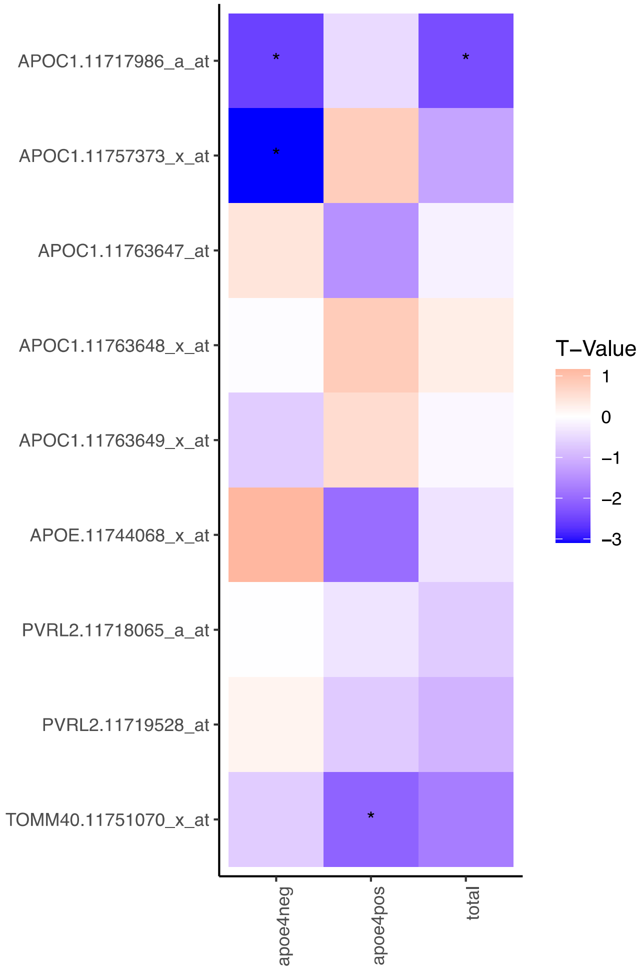

**Supplementary Figure 3: Significant differentially (a) AD and (b) APOE methylated networks in Brain**

**a**

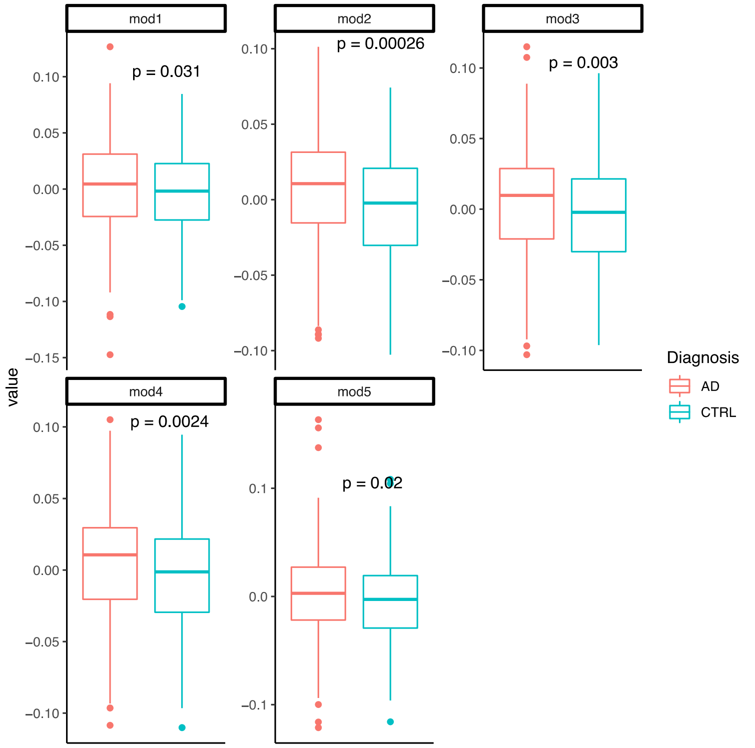


**b**

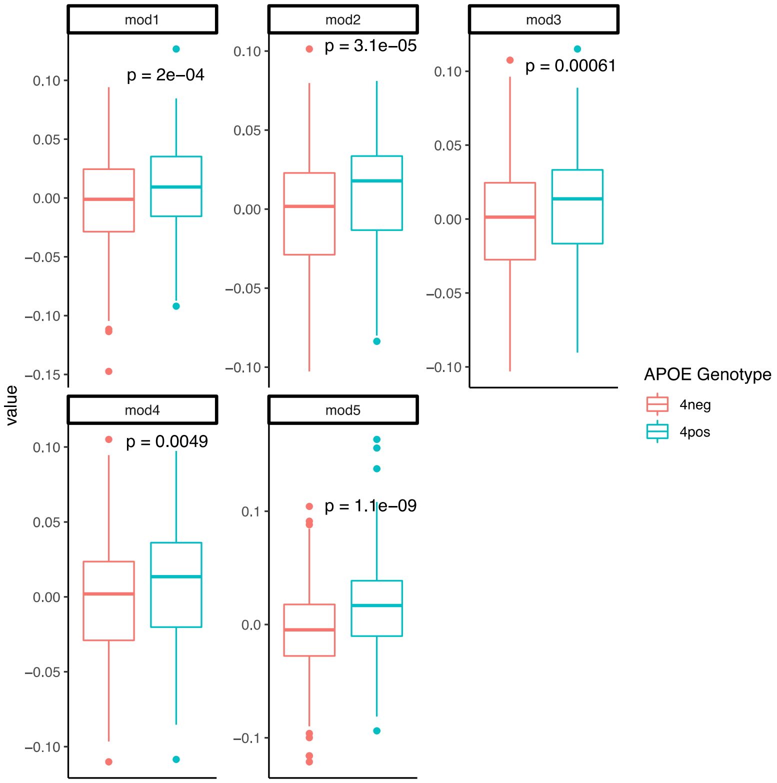

**Supplementary Figure 4: Significant differentially (a) AD and (b) APOE methylated networks in Blood**

**a**

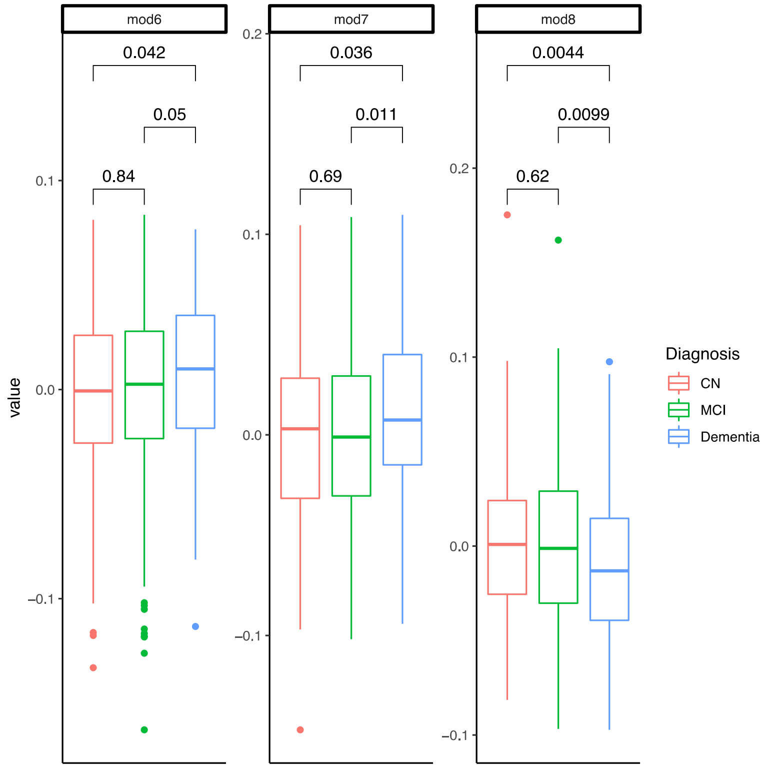


**b**

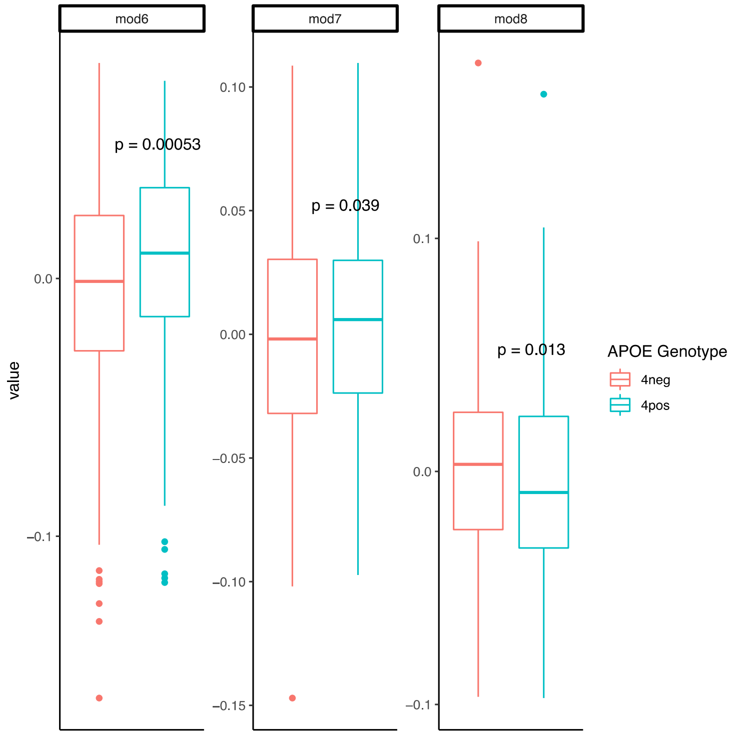
